# Privacy-preserving AUC Computation in Distributed Machine Learning with PHT-meDIC

**DOI:** 10.1101/2025.01.14.25320558

**Authors:** Marius de Arruda Botelho Herr, Cem Ata Baykara, Ali Burak Ünal, Nico Pfeifer, Mete Akgün

**Affiliations:** Methods in Medical Informatics, Department of Computer Science, University of Tübingen, Sand 14, Tübingen 72076, Germany; Medical Data Privacy and Privacy-Preserving ML on Healthcare Data, Department of Computer Science, University of Tübingen, Sand 14, Tübingen 72076, Germany; University Hospital Tübingen, Institute for Translational Bioinformatics, Tübingen, 72072, Germany

## Abstract

Ensuring privacy in distributed machine learning while computing the Area Under the Curve (AUC) is a significant challenge because pooling sensitive test data is often not allowed. Although cryptographic methods can address some of these concerns, they may compromise either scalability or accuracy. In this paper, we present two privacy-preserving solutions for secure AUC computation across multiple institutions: (1) an exact global AUC method that handles ties in prediction scores and scales linearly with the number of samples, and (2) an approximation method that substantially reduces runtime while maintaining acceptable accuracy. Our protocols leverage a combination of homomorphic encryption (modified Paillier), symmetric and asymmetric cryptography, and randomized encoding to preserve the confidentiality of true labels and model predictions. We integrate these methods into the Personal Health Train (PHT)-meDIC platform, a distributed machine learning environment designed for healthcare, to demonstrate their correctness and feasibility. Results using both real-world and synthetic datasets confirm the accuracy of our approach: the exact method computes the true AUC without revealing private inputs, and the approximation provides a balanced trade-off between computational efficiency and precision. All relevant code is publicly available at https://github.com/PHT-meDIC/PP-AUC, facilitating straightforward adoption and further development within broader distributed learning ecosystems.

**Author summary:** A commonly used metric to evaluate the performance of machine learning models is the Area Under the Curve (AUC). Calculating the AUC in distributed machine learning settings is challenging because data cannot be shared between institutions due to privacy concerns. To address this, we developed two privacy-preserving methods: one that calculates the exact AUC securely and another that provides faster approximations with high accuracy. These methods use advanced encryption techniques to protect sensitive data while enabling secure collaboration. We tested them in a real-world healthcare platform called PHT-meDIC and demonstrated their effectiveness. The code is publicly available at https://github.com/PHT-meDIC/PP-AUC to support wider adoption.

## Introduction

The Area Under the Curve (AUC) is one of the most widely used metrics for evaluating the performance of binary classifiers, providing a robust summary of a model’s ability to distinguish between positive and negative classes. In centralized machine learning settings, computing AUC is straightforward as both prediction scores and true labels are readily accessible. However, in distributed machine learning frameworks, such as federated learning [1–3], ensuring the privacy of participants’ data, particularly true labels, becomes a critical challenge. Labels often contain privacy-sensitive information, and sharing them across multiple parties in a distributed environment could lead to privacy breaches.

Most existing approaches to compute the AUC in distributed settings privacy-preserving rely on differential privacy (DP) mechanisms [4–6], which introduce noise to protect sensitive information but yield approximate AUC values. Other methods, such as ROC-GLM [7] in DataSHIELD [8], use statistical techniques but carry overhead and remain constrained to specific software ecosystems. Cryptographic solutions based on multi-party computation (MPC) [9] enable exact AUC calculation but frequently suffer from scalability issues or significant communication overheads.

This paper presents PP-AUC, two novel cryptographic methods for distributed, privacy-preserving exact (DPPE) and approximation (DPPA) AUC computation that addresses these limitations. Our exact method combines cloud-based Paillier homomorphic encryption with symmetric and asymmetric cryptography and randomized encoding to enable AUC’s secure and exact computation in distributed environments without revealing sensitive data. The approximation method utilizes similar technologies but only approximates the AUC based on a predefined number of specific thresholds used to calculate the AUC.

Unlike existing methods, DPPE-AUC can compute the global AUC across any number of participants and handle tie conditions, where multiple samples may have identical prediction scores while ensuring both accuracy and scalability. The DPPA-AUC approach addresses the computational inefficiencies by providing an approximate AUC computation using full homomorphic secure aggregation. However, it introduces deviations in accuracy similar to differential privacy methods.

We integrate DPPE- and DPPA-AUC methods within the PHT-meDIC platform [10], a distributed learning platform tailored for privacy-preserving analysis of healthcare data. To evaluate the performance of both methods, we conduct experiments on real-world healthcare datasets and synthetic data, demonstrating that our protocol achieves exact AUC computation while scaling linearly with the number of samples. All runs of the DPPE-method are compared in terms of runtime and correctness to the DPPA-AUC method. Our implementation is publicly available in Python and can be easily adapted to other distributed machine-learning systems. In the PHT-meDIC setting, the analogy of stations and trains represents data stations acting as secure endpoints where algorithms (trains) travel to access and process data locally, ensuring privacy while enabling collaborative computations.

## Background

### Area Under Curve

There are several approaches to summarize the plot-based evaluation metrics for machine learning models. One of the most popular and effective ones is AUC. It summarizes the performance evaluation of the model by giving the area under the curve of the utilized plot-based evaluation metric. This paper employs the receiver operating characteristic (ROC) curve as the plot-based evaluation metric. The ROC curve aims to evaluate the performance of a model by considering the true positive rate sensitivity and the model’s specificity. It plots the false positive rate (FPR) on the x-axis and the true positive rate (TPR) on the y-axis. To summarize the evaluation of the ROC curve, AUC computes the area under the ROC (AUROC) via the following formula:

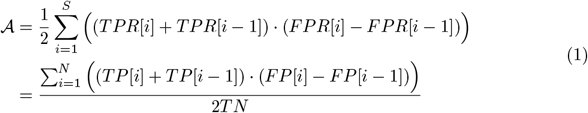

Where *S* is the set of samples whose prediction scores are different than their subsequent sample’s prediction score, 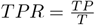 is the true positive rate computed by dividing the number of true positive, that is *TP*, by the total number of positive samples, that is *T*, and, similarly, 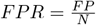 is false positive rate computed by dividing the number of false positives, that is *FP*, by the number of negative samples, that is *N*. Equation 1 can compute the exact AUC whether the underlying prediction scores have a tie condition, where the tie condition indicates that at least two samples have the same prediction score.

### Modified Paillier Cryptosystem

The modified Paillier system [11, 12] supports the addition of two ciphertexts and the multiplication of a ciphertext with a plaintext constant. This allows users to perform computations on ciphertexts, which results in an identical output as that performed on plaintexts. In this system, the public key is (*n, g, h* = *g*^*x*^). *n* is the product of two safe primes: *z* and *y*. The secret key is 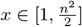 and *g* that is in order of 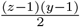 equals to −*a*^2*n*^ where 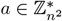

- **Encryption:** The message *m* ∈ 𝕫_*n*_ is encrypted as follows: *c*_1_ = *g*^*r*^ mod *n*^2^ and *c*_2_ = *h*^*r*^(1 + *mn*) mod *n*^2^ where a random *r* ∈ [1, *n/*4].
- **Decryption:** To recover *m*, the decryption of (*c*_1_, *c*_2_) is performed as follows: 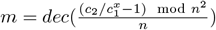
- **Proxy Re-encryption:** The secret key *x* is split into two shares such that *x* = *x*_1_ + *x*_2_. In the modified Paillier system the ciphertext (*c*_1_, *c*_2_) can be partially decrypted to 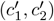 as follows: 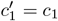 and 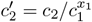 mod *n*^2^ Then, 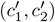 can be decrypted to *m* using *x*_2_ instead of *x* with the decryption method described above.

### Homomorphic Properties

The modified Paillier cryptosystem is homomorphic with respect to both addition and scalar multiplication. This means that certain operations on encrypted data correspond directly to the same operations on the underlying plaintexts without requiring the data to be decrypted first. Such homomorphic properties are fundamental in enabling secure computations on encrypted data.

- **Addition**: Let (*c*_1,1_, *c*_1,2_) be the encryption of a message *m*_1_ and (*c*_2,1_, *c*_2,2_) be the encryption of a message *m*_2_. In the modified Paillier scheme, “adding” these ciphertexts (through the group operation in 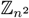, typically component-wise multiplication) produces a new ciphertext (*c*_3,1_, *c*_3,2_). (*c*_3,1_, *c*_3,2_) = (*c*_1,1_· *c*_2,1_ mod *n*^2^, *c*_1,2_· *c*_2,2_ mod *n*^2^). Decrypting (*c*_3,1_, *c*_3,2_) yields *m*_1_ + *m*_2_ mod *n*. Hence, dec(*c*_3,1_, *c*_3,2_) = *m*_1_ + *m*_2_ (mod *n*). This property is crucial in scenarios where multiple values must be summed securely.
- **Scalar Multiplication**: Additionally to addition, the Paillier cryptosystem also supports the multiplication of an encrypted value by a plaintext constant. Suppose (*c*_1_, *c*_2_) encrypts a message *m*, and let *k* be a known integer in 𝕫_*n*_. Then raising the ciphertext to the power *k* corresponds to multiplying the underlying plaintext by *k*:

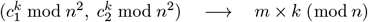 Consequently, dec 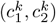 This ability to apply scalar multiplication homomorphically is useful for computing weighted sums or scaling factors while preserving the confidentiality of the underlying data.

These homomorphic properties allow us to calculate the exact AUC with partial decryption in proxy re-encryption settings — without ever exposing the underlying plaintexts.

### Randomized Encoding for Multiplication

One of the privacy-enhancing techniques that we use in this study is Randomized Encoding (RE) [13, 14]. The idea of RE is to use random values to hide a function’s inputs and reveal only its output. It creates components of encoding of the desired function by using random values. Then, the output of this function can only be obtained by combining these components in a certain way. There are REs for several functions in the literature for privacy-preserving machine learning [15, 16]. Among those, we benefit from the RE for *multiplication* [17].

**Definition 1 (Modified RE for Multiplication [17])** *Let f be a function of x*_1_ *and x*_2_ *defined over a ring* R *such that f* (*x*_1_, *x*_2_) = *x*_1_ *x*_2_. *f can be perfectly encoded by the function* 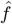 *defined as follows:*

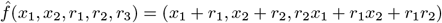

*where r*_1_, *r*_2_ *and r*_3_ *are uniformly chosen random values. In order to obtain the result of y* = *f* (*x*_1_, *x*_2_), *one needs to compute c*_1_· *c*_2_ − *c*_3_ *given the encoding* (*c*_1_, *c*_2_, *c*_3_).

### PHT-meDIC

A prominent representative of distributed analytics is the Personal Health Train (PHT), created under the GO FAIR initiative [18] to enable privacy-preserving data analysis on medical data collected from hospitals and research projects. It was further developed by the German National Chapter through standards, guidelines, and reference implementations [19]. Since 2019, different PHT ecosystems have emerged within the German medical informatics initiative (MII) [20]. Within MII, the PHT-meDIC is one technology for distributed analysis between data integration centers (DIC), which securely host input data. PHT-meDIC [10] is a distributed learning platform developed for privacy-preserving healthcare data analysis within the MII. Built around an ‘algorithm-to-data’ paradigm, it ensures that sensitive patient information remains under local control at each institution, eliminating the need for data pooling or replication. The platform’s container-based approach facilitates the secure execution of complex analytics while reducing infrastructure modifications at local hospital sites. Furthermore, the PHT-meDIC is interoperable [21] with other DIC distributed learning platforms such as PADME [22]. PHT-meDIC employs data encryption at rest and follows an honest but curious threat model to safeguard sensitive healthcare data. Moreover, it signs each algorithm and maintains a chain of digital signatures, ensuring no manipulation throughout the distributed analysis process. In addition, PHT-meDIC provides built-in governance mechanisms,such as mandatory code review and approval workflows, to uphold compliance and maintain trust among participating institutions. The key generation within the PHT-meDIC platform can be directly handled by the user within the desktop app for symmetric encryption protocols (RSA) and Paillier keys or, as in our case, within an initialization station. This station does not contribute data to the analysis but handles tasks like additional key creation (Cloud-Paillier key components).

### Related Work

DP has become a popular framework for protecting sensitive data in federated analysis, as it offers well-defined privacy guarantees by adding controlled noise [23]. However, DP approaches typically compute approximate metrics, leading to discrepancies between a model’s theoretical and actual performance. Besides the approximation approach, DP method [24] assumes the proxy server is honest-but-curious and local or global input can be obtained. Some recent efforts explore hybrid methods that combine DP with secure aggregation or limited cryptographic primitives to improve accuracy [25]. However, these hybrid strategies can still introduce computational and communication overhead

Statistical models like ROC-GLM [7], implemented within the DataSHIELD framework, enable distributed AUC estimation but have platform-specific dependencies and may not scale seamlessly to large datasets. In contrast, multi-party computation (MPC) protocols [9] facilitate exact computation by performing secure operations across multiple sites. Although MPC ensures that raw data remain private, it often involves complex implementation details and high communication costs. Consequently, advanced MPC systems — such as those based on homomorphic encryption — are still not widely used for large-scale AUC estimation. These constraints highlight the need for methods that maintain exactness, reduce overhead, and scale effectively across multiple sites without sacrificing privacy guarantees.

## Materials and Methods

### Preliminaries of the Protocol

Assume we have *n* participating stations (S_1_, S_2_, …, S_n_) and a initialization and proxy station S_0_ and S_P_ in the network. The initialization station S_0_ and the proxy station S_P_ do not participate in the analysis but act as an intermediary for key creation and secure computations. Each station holds its local data, and the machine learning model has already produced prediction values and randomly assigned flag subjects for privacy preservation.

Let *M* be the number of samples, and let the masked prediction values at each station be denoted as {*pre*}, where the predictions are masked as {*pre*} = (*r*_1_· *pre*) + *r*_2_, with *r*_1_ and *r*_2_ being randomly generated values to obscure the real prediction data.

Our protocols utilize the following functions: asymmetric encryption, symmetric encryption, symmetric key generation, Paillier encryption, and random number generation. These functions are defined as follows:

- RANDS(): Generates random integers.
- SYMM(): Generates a symmetric Fernet key for a station *S*_*i*_.
- ENC(*M, K*): Encrypts a message *M* using a key *K*, which can be an RSA, Paillier, or symmetric key.
- DEC(*C, K*): Decrypts a ciphertext *C* using a key *K*, which can be an RSA, Paillier, or symmetric key.
- ADD(*C*_1_, *C*_2_): A homomorphic function that performs the addition of ciphertexts *C*_1_ and *C*_2_, encrypted with the same public key *PK*.
- ADD(*C, M*): A homomorphic function that performs the addition of a ciphertext *C* and a plaintext *M*. The plaintext *M* is first encrypted using a public key *PK*.
- MUL(*C, M*): A Paillier function that multiplies a plaintext *M* with a ciphertext *C* under a public key *PK*.

### DPPE-AUC

In this section, we introduce our distributed privacy-preserving method for exact AUC computation, referred to as DPPE-AUC, which has been seamlessly integrated into the PHT-meDIC platform.

#### Initialization

Each station is equipped with an RSA public and private key pair 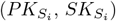. The RSA public key of every station is accessible to all other stations within the network. Station S_0_ generates a public and private key pair (*PK*_*P*_, *SK*_*S*_) for a modified Paillier cryptosystem. The private key *SK*_*S*_ is randomly divided into two parts,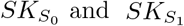, such that 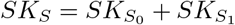.

The public key *PK*_*P*_ is distributed to all stations. The split private key 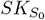 is securely transmitted to the proxy station S_P_, while 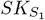 is securely sent to each station S_i_, where *i* ∈ {1, 2, …, *n*}. The distribution of these keys is performed securely by encrypting them with the RSA public keys of the respective stations.

This initialization process is carried out using a train (image) updated by station S_0_, which sequentially visits all stations along the route (S_1_, S_2_, …, S_n_, S_P_).

#### Client Computation

During the execution of Algorithm 1, each station generates a symmetric key *k*_*i*_. The first station generates a random value *r*_1_, used to obscure the prediction values by multiplication. At each station, an additional random component *r*_2_ is calculated as *r*_2_ = (*pre* mod *r*_1_), further masking the prediction values. These masked predictions are encrypted using the symmetric key *k*_*i*_, which is encrypted with the proxy station’s public key 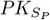.

The random value *r*_1_ is encrypted by the RSA public keys of stations other than S_0_ and S_P_. Additionally, binary labels and flag values ∈ {0, 1} are encrypted using Paillier public key *PK*_*P*_. The encrypted results are stored within the train image and passed to the next station in the route. The subsequent stations decrypt ⟨*r*_1_⟩ using their RSA private key. The process of obfuscating prediction values and encrypting labels and flags continues analogously across all stations.

##### Algorithm 1

*dppe-auc* Station Protocol - Step I

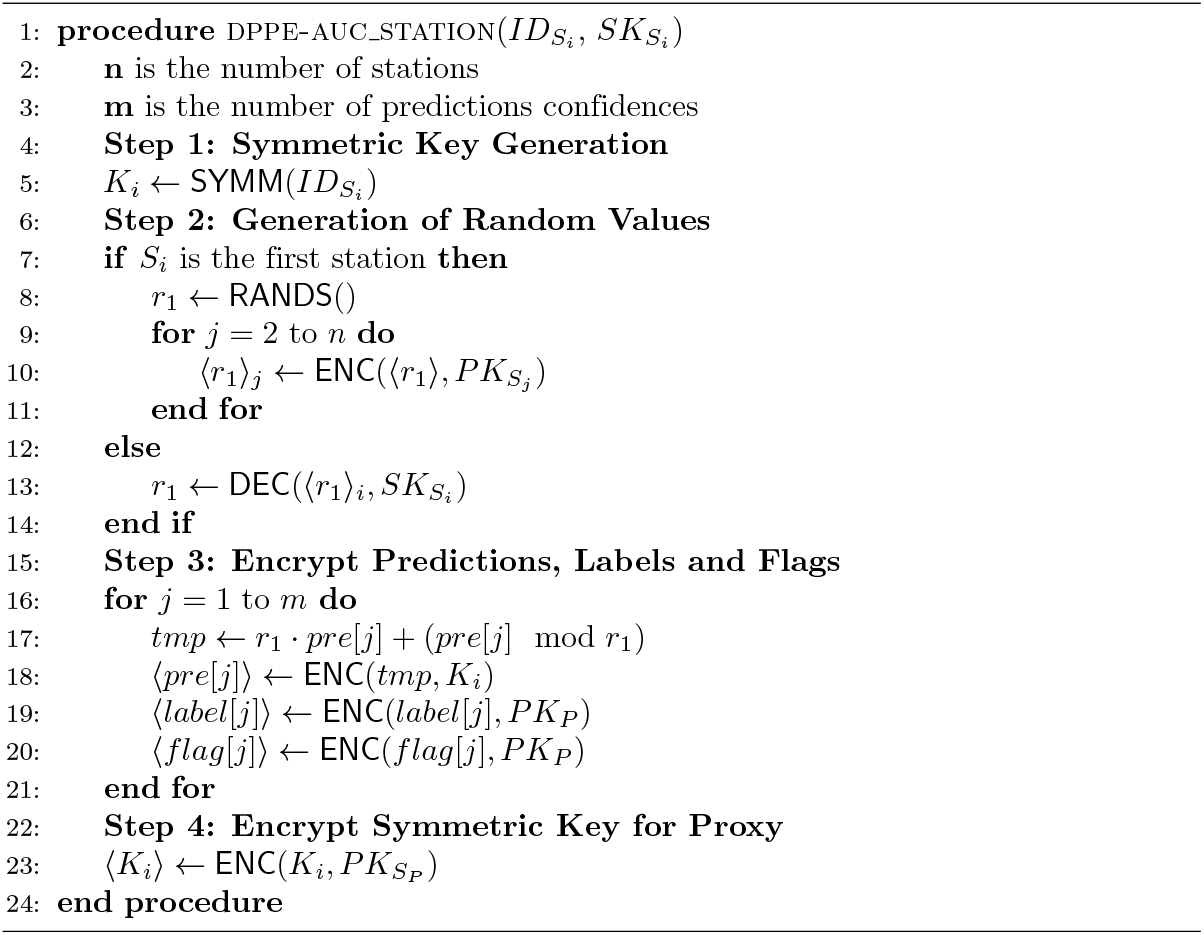

#### Proxy Computation

The proxy station retrieves the train image and executes Algorithm 2. First, it decrypts the symmetric keys (⟨*K*_1_⟩, ⟨*K*_2_⟩, …, ⟨*K*_*n*_⟩) using its private asymmetric key 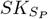 The masked prediction values from all stations are decrypted and concatenated into a single table along with the corresponding encrypted labels and flags. *S*_*P*_ then sorts this table according to the masked prediction confidences while keeping the actual prediction values hidden, ensuring that it does not access the real input data.

Based on the Paillier-encrypted labels and flag values, the proxy computes the encrypted true positive ⟨*TP*⟩ and false positive ⟨*FP*⟩ values. For each masked prediction value *pre*[*i*], the proxy sums all labels greater than *pre*[*i*] to calculate ⟨*TP* [*i*] ⟩. The corresponding ⟨*FP* [*i*]⟩ value is determined by subtracting ⟨*TP* ⟩ [*i*] from the sum of the flag values greater than *pre*[*i*].

Due to the limitations of Paillier encryption, the proxy cannot directly perform multiplications on encrypted values. Instead, it delegates these computations to the stations. However, to prevent stations from inferring private information about other participants, we apply randomized encoding techniques to mask both the numerator (*N*) and denominator (*D*) values.

For the denominator *D*, the proxy computes the sum of all labels as ⟨*TP*_*A*_⟩ and the sum of all flag values minus ⟨*TP*_*A*_⟩ (Equation 2) as ⟨*FP*_*A*_⟩. These values are multiplied by randomly generated constants *a* and *b*, respectively, using Paillier multiplication.

The results are then used to compute the randomized encoding components of the denominator: ⟨*D*_1_⟩, ⟨*D*_2_⟩, and ⟨*D*_3_⟩, with random integers 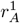 and 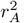:

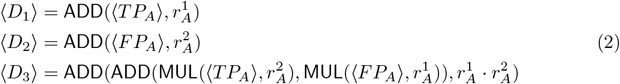

For the numerator *N*, the proxy computes the difference in successive false positive values as ⟨*dFP* [*i*] ⟩ = ⟨*FP* [*i*] ⟩ − ⟨*FP* [*i* − 1] ⟩ and the sum of successive true positive values ⟨*sTP* [*i*] ⟩ = ⟨*TP* [*i*] ⟩ + ⟨*TP* [*i* − 1] ⟩ across all threshold values (Equation 3). All ⟨*sTP* [*i*] ⟩ and ⟨*dFP* [*i*] ⟩ values are multiplied by *a* and *b*, respectively. To handle ties in prediction values, we use the method from [9] to compute unique indices for the masked predictions. The proxy generates three random values 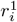,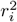 and *z*_*i*_ for each index *i*. All *z*_*i*_ values are summed to zero. These random values are used to compute the randomized encoding components for the numerator:

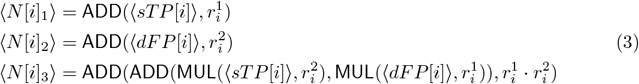

To improve runtime efficiency, the sum of all ⟨*N* [*i*]_3_⟩ values is computed as ⟨*N*_3_⟩. The proxy station then partially decrypts all random components ⟨*D*_1_⟩, ⟨*D*_2_⟩, ⟨*D*_3_⟩, ⟨*N* [*i*]_1_⟩, ⟨*N* [*i*]_2_⟩, and ⟨*N*_3_⟩, where *i* ∈ {1, 2, …, *m*}, using its partial private key 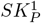. These components are written to the train results, and the proxy pushes the updated image back to the stations.

#### Final Computation

Each station S_i_ pulls the updated image and extracts the partially decrypted random components and fully decrypts them using its private key *SK*_1_. Station S_i_ can then locally compute the DPPE-AUC using the following equation:

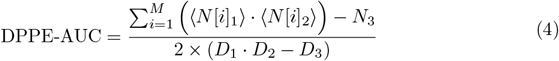

It is impossible to eliminate the randomness present in the fully decrypted values for the stations. The only method to completely remove all randomness from these values is to calculate the AUC using Equation above.

### DPPA-AUC

In this section, we provide a detailed explanation of our approximate AUC computation method, referred to as DPPA-AUC.

The use of approximate AUC for privacy-preserving model performance evaluation in distributed settings was first proposed by Sun et al. [24]. Sun et al. employed differential privacy to compute the AUC score of a global model in a federated environment. Although DPPA-AUC adopts the same underlying approximation technique, rather than differential privacy, it uses Paillier cryptosystem and randomized encoding to compute the approximate AUC value within the PHT-meDIC platform.

#### Algorithm 2

*dppe-auc* Aggregation Protocol

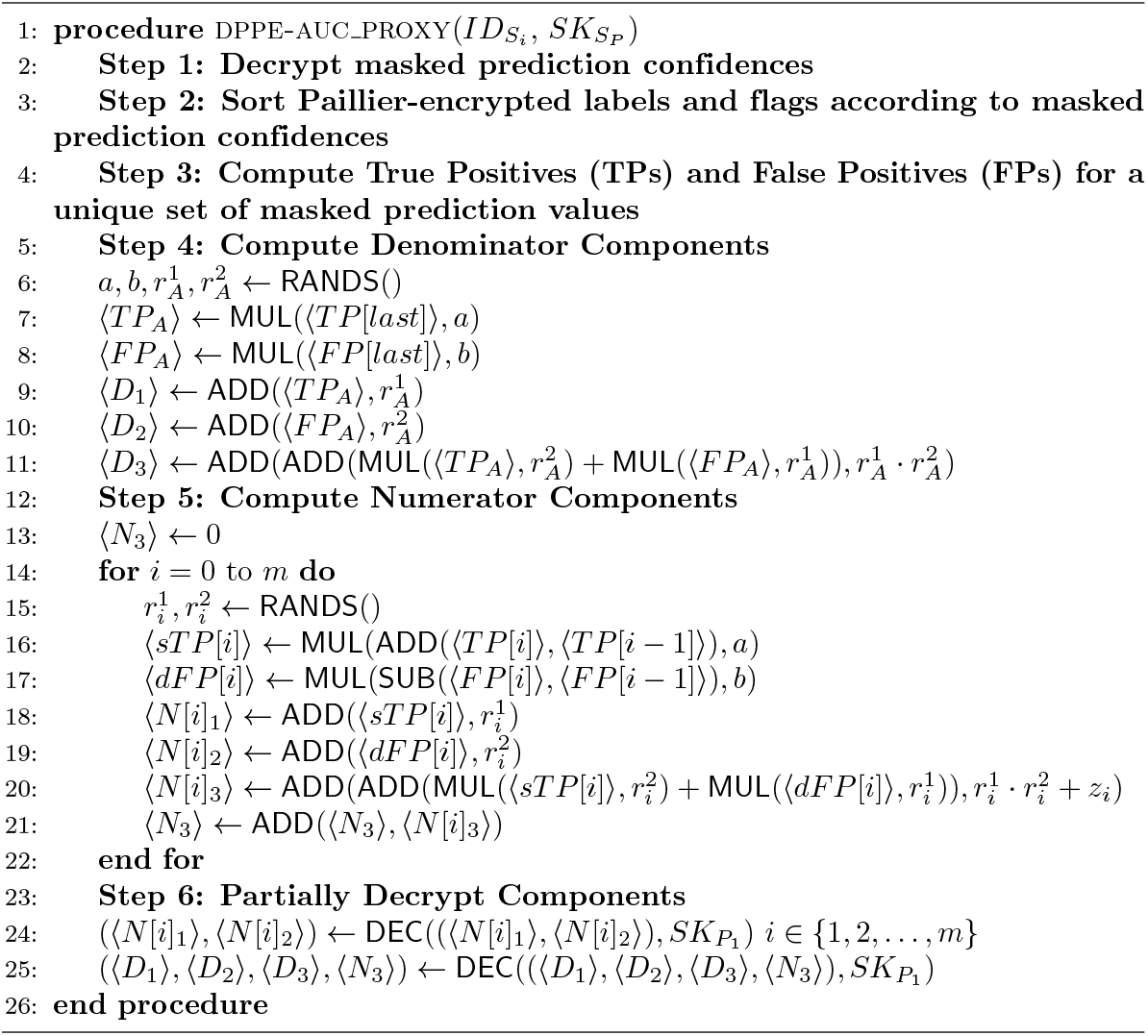

#### Initialization

The initialization phase of DPPA-AUC is identical to the DPPE-AUC. Each station is equipped with an RSA public and private key pair (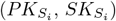), and the RSA public key of every station is accessible to all others. Station *S*_0_ generates a modified Paillier cryptosystem key pair (*PK*_*P*_, *SK*_*S*_) and splits *SK*_*S*_ into 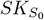, distributing these securely via RSA encryption. The public key *PK*_*P*_ is shared across the network, while the private key parts are distributed along a predefined route using a train (image) updated by S_0_.

#### Client Computation

During the execution of Algorithm 3, each station computes their TP and FP for a set of predefined decision points *D*. The number of prediction confidences is denoted as *m*.

For each decision point *D*[*i*], the algorithm accumulates the counts of *ones* (representing true labels equal to 1) and *zeros* (representing true labels equal to 0). These counts are updated while traversing the prediction values until the confidence values fall below the current decision point *D*[*i*]. If the last prediction value is reached and has not yet been visited, the algorithm ensures it is processed to finalize the counts.

The computed true positives *TP* [*i*] and false positives *FP* [*i*] for each decision point are then encrypted using the Paillier public key *PK*_*P*_. The encrypted values ⟨*TP* [*i*]⟩ and ⟨*FP* [*i*]⟩ are passed to the next station or the proxy for further processing.

##### Algorithm 3

*dppa-auc* Station Protocol

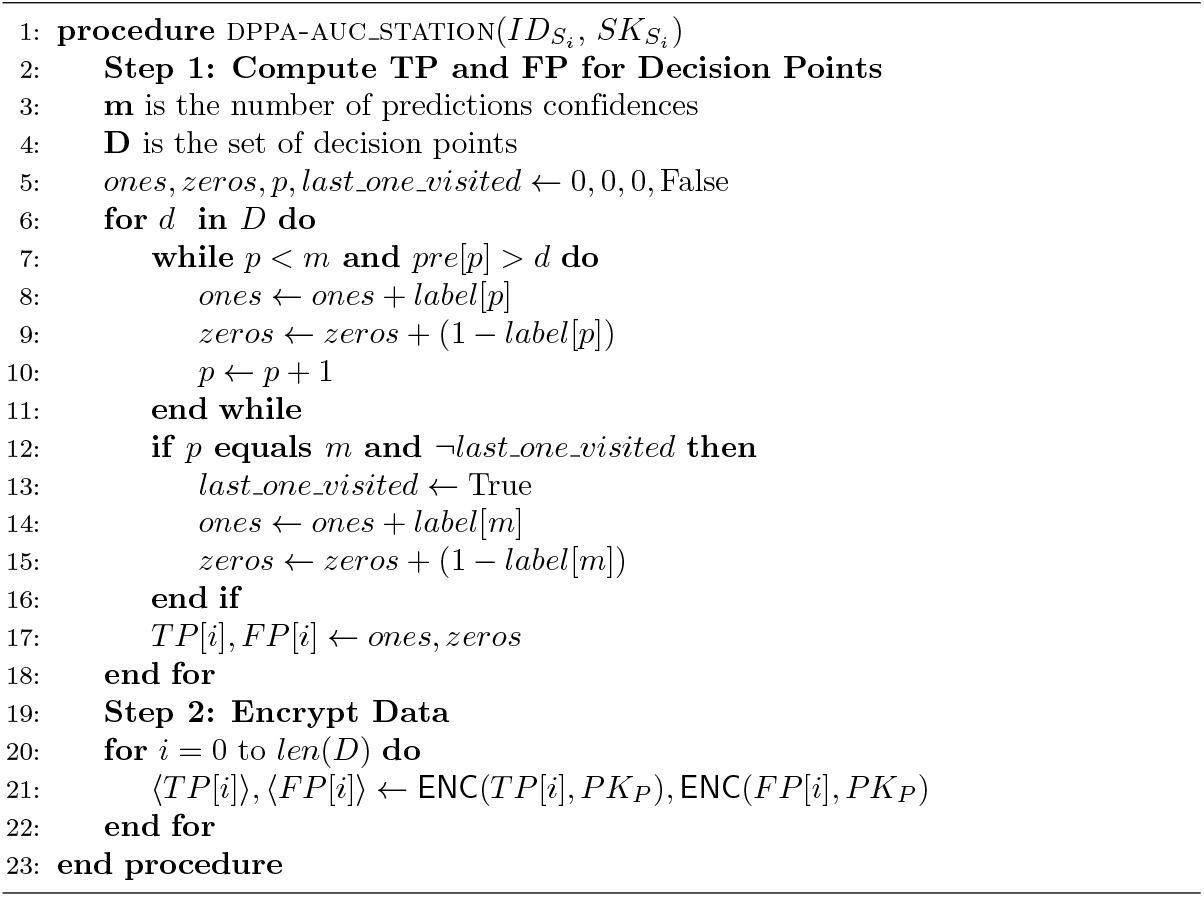

#### Proxy Computation

During the execution of Algorithm 4, the proxy station aggregates the TP and FP values across all stations for a set of decision points *D*. For each decision point *D*[*j*], the proxy computes the global sums ⟨*TP*_*global*_[*j*]⟩ and ⟨*FP*_*global*_[*j*]⟩ by adding the encrypted contributions 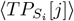 and 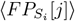 from all stations using the Paillier encryption scheme.

Next, the proxy calculates the final TP and FP values. For each decision point *j* (from the second point onward), the proxy adds the cumulative values from the previous decision point to the current point for ⟨*TP*_*final*_[*j*] ⟩. For ⟨*FP*_*final*_[*j*] ⟩, the proxy computes the difference between the current and previous FP values. These calculations are performed securely using Paillier operations to preserve data privacy.

From this point on, Algorithm 4 follows the same steps as Algorithm 2 for computing the numerator and denominator components, as well as partially decrypting these components. Specifically, the proxy computes randomized encoding components for the numerator and denominator using securely scaled true positive and false positive values. The proxy then partially decrypts these randomized components using its partial private key 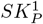 and shares them back with the stations.

##### Algorithm 4

*dppa-auc* Aggregation Protocol

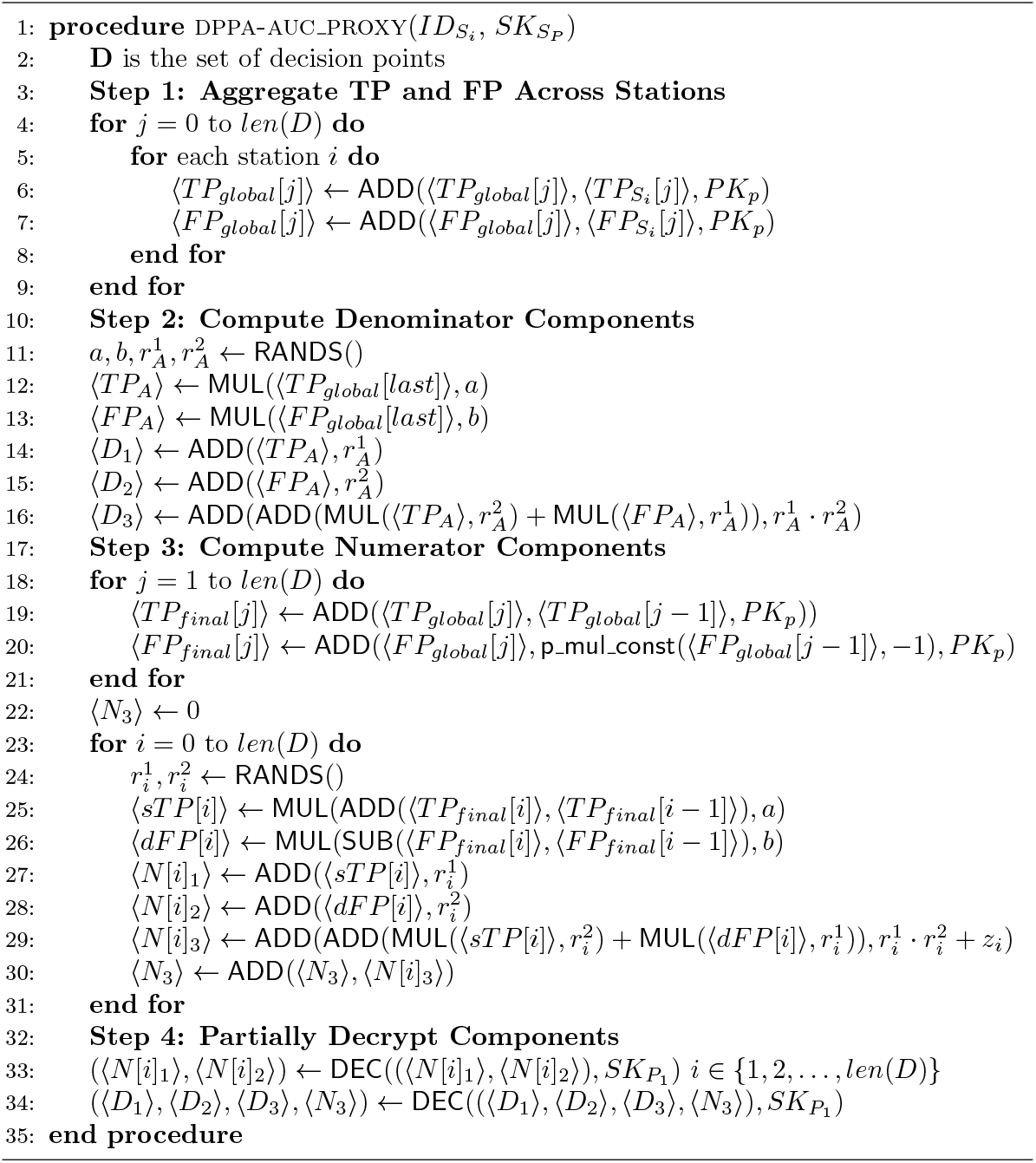

#### Final Computation

DPPA-AUC computation utilizes the same Equation 4 as the DPPE-AUC to compute the final AUC value locally at each station.

### Security Analysis

Assume there are *N* stations in the PHT framework. An adversary controlling (*N* − *M*) stations cannot infer any information about the honest *M* stations’ predictions, labels, or flags. Labels and flags are encrypted using the modified Paillier cryptosystem, with encryption performed using the proxy station’s public key. Since stations only hold a share of the private key of the proxy, an adversary controlling *N* − *M* stations cannot access the other part of the private key. Predictions are encrypted with AES-GCM, and the symmetric key is encrypted using the proxy station’s RSA public key. Consequently, without the proxy’s private RSA key, the adversary cannot access the predictions of honest stations.

An adversary controlling only the proxy station cannot access private data, as labels and flags are encrypted with the proxy’s public key. The proxy station cannot decrypt this information since it only knows a portion of its private key. Although predictions are masked with two random values, the adversary can observe the masked values but cannot infer meaningful differences between them due to the presence of dummy values. These dummy values significantly reduce the likelihood of identifying the real samples, preventing the adversary from confidently estimating the relative differences between actual prediction values.

Additionally, the adversary cannot determine the number of predictions at each station due to inserting dummy values. However, if an adversary controls both the proxy station and at least one participating station, they could decrypt all predictions, labels, and flags by combining the private keys. For this reason, our security model assumes no collusion between the proxy station and other stations.

The security of our scheme depends on the security properties of the modified Paillier system and randomized encoding. The security of the Paillier cryptosystem was analyzed by [11], which showed two important guarantees: one-wayness, meaning that the ciphertext and public key cannot be used to deduce the plaintext, and semantic security, ensuring that an adversary cannot match plaintext-ciphertext pairs even when given two plaintexts. The definitions and security proofs for the randomized encoding of addition and multiplication used in this scheme are provided by [17].

This security analysis applies to both DPPE-AUC and DPPA-AUC, since the only methodological difference (exact vs. approximate thresholds) does not affect the underlying cryptographic primitives or trust assumptions.

### Showcase and Experiments

To evaluate the efficacy and privacy-preserving capabilities of our DPPE- and DPPA-AUC methods, we conducted experiments using both real-world HIV-1 V3-loop sequence data and synthetic datasets, benchmarking against ground truth (GT) AUC using the *roc auc score* function from the scikit-learn metrics library [26].

#### Showcase: HIV-1 Coreceptor Binding Prediction

For the primary showcase, we utilized 10462 HIV-1 V3-loop sequences annotated with coreceptor binding data [27]. The binary classification task is to predict binding preferences for coreceptors *CCR*5 and *CXCR*4, with data distributed across three stations for decentralized analysis. Each station trained a local model using its partitioned dataset, while a 10% holdout set was maintained for independent evaluation. This setup allowed us to test our method’s effectiveness in preserving data privacy across distributed sites.

For performance reasons, we employed a flag patient generation approach, injecting synthetic samples only within value range of actual patient data to not increase the number of thresholds. We performed all experiments and showcases on a local computer (Apple® Mac mini, M4Pro, 48GB memory).

We computed the corresponding DPPE- and DPPA-AUC components for our method and derived the difference to the GT. Our experiments included the DPPE- and DPPA-AUC method compared in terms of time and deviations from GT AUC score. The results in Table 1 highlight a trade-off between accuracy and computational efficiency: DPPE-AUC achieves near-perfect accuracy with a minimal difference from the GT but incurs a higher computational cost, while DPPA-AUC demonstrates faster runtime with a slightly larger but acceptable deviation from the GT AUC.

**Table 1.** Comparison of DPPE-AUC and DPPA-AUC methods based on runtime, the difference from the ground truth AUC, and the number of thresholds used.

| Method | Time (s) | Deviation to GT | Thresholds |
| --- | --- | --- | --- |
| DPPE-AUC | 38.71 | 1.11E-16 | 145 |
| DPPA-AUC | 26.499 | 0.0037 | 100 |

For each run, Across ten runs, the average difference between the GT and DPPE-AUC values was 10^*−*16^, likely attributable to floating-point arithmetic precision limits in Python.

#### Experiments

In the showcase described above, we observed a high AUC score, which resulted in numerous repetitive prediction values. This led to data bias, making a performance evaluation based on increasing sample size and station count impractical. To address this, we generated synthetic data to examine the impact of these values more accurately. At each station, subjects were randomly assigned prediction values between 0 and 1 and labels from the set 0, 1. Random flag patients were created identically to those in the showcase. We compared the GT AUC with the DPPE- and DPPA-AUC, excluding flag samples in the GT calculation to validate our method. Across experiments, differences between GT and DPPE-AUC values remained consistently at 10^*−*18^.

#### Experiment 1

In this experiment, we analyzed the runtime performance of both methods compared to GT AUC. The total dataset size was fixed at 1500 subjects, combining both actual and dummy (flag) patients. The experiment aimed to simulate a realistic PHT-meDIC environment, where an increasing number of participating stations (or input parties) operate decentralized. As the number of sites increased, the sample size per site was proportionally reduced per site.

This configuration allowed us to analyze how computational efficiency scales with the number of sites while maintaining a constant total sample size. Each site independently computed its local AUC contribution iteratively, with the overall execution time determined by the combined decentralized encryption procedures. The synthetic data generation in this experiment mimicked a real-world scenario by introducing biases around prediction values near 0 and 1. Flag data was generated only for existing subjects to obscure the data distribution, ensuring no increase in the number of threshold values. This approach effectively minimized computational overhead while preserving data realism.

As expected, the runtime increased with the number of participating sites in Fig 3 due to decentralized computations overhead. Both methods demonstrated robust performance, though DPPE-AUC required slightly more time than DPPA-AUC due to its complete processing of input data, while DPPA-AUC only uses 100 approximation points.

**Fig 1.**
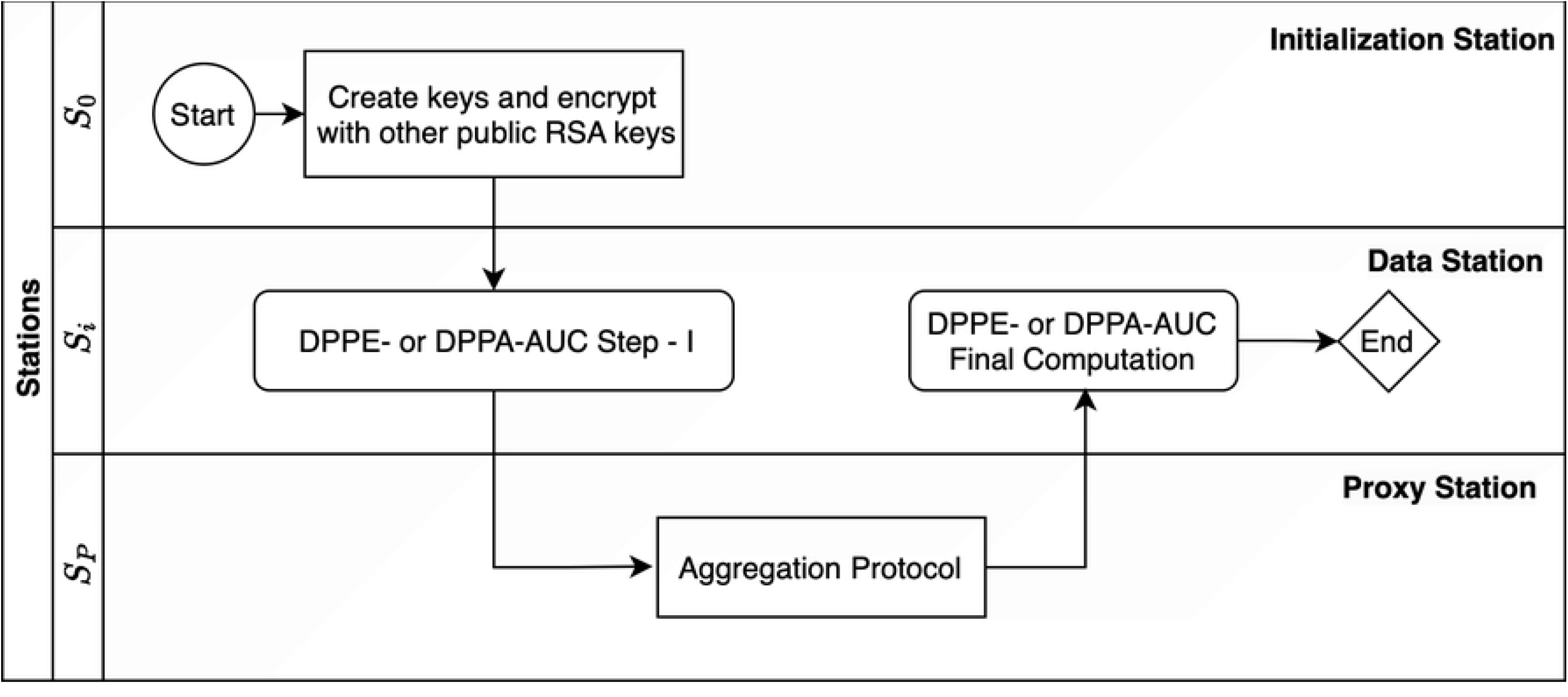
Overview of the DPPE- and DPPA-AUC Process: The workflow involves three types of stations: the initialization Station (S_0_), Data Stations (S_i_), and Proxy Station (S_P_). The process starts at S_0_ with Paillier key generation and encryption using RSA public keys. S_i_ perform DPPE- or DPPA-AUC Step I, followed by an Aggregation Protocol at the S_P_. The results are then processed in final computation at the S_i_, completing the protocol.

**Fig 2.**
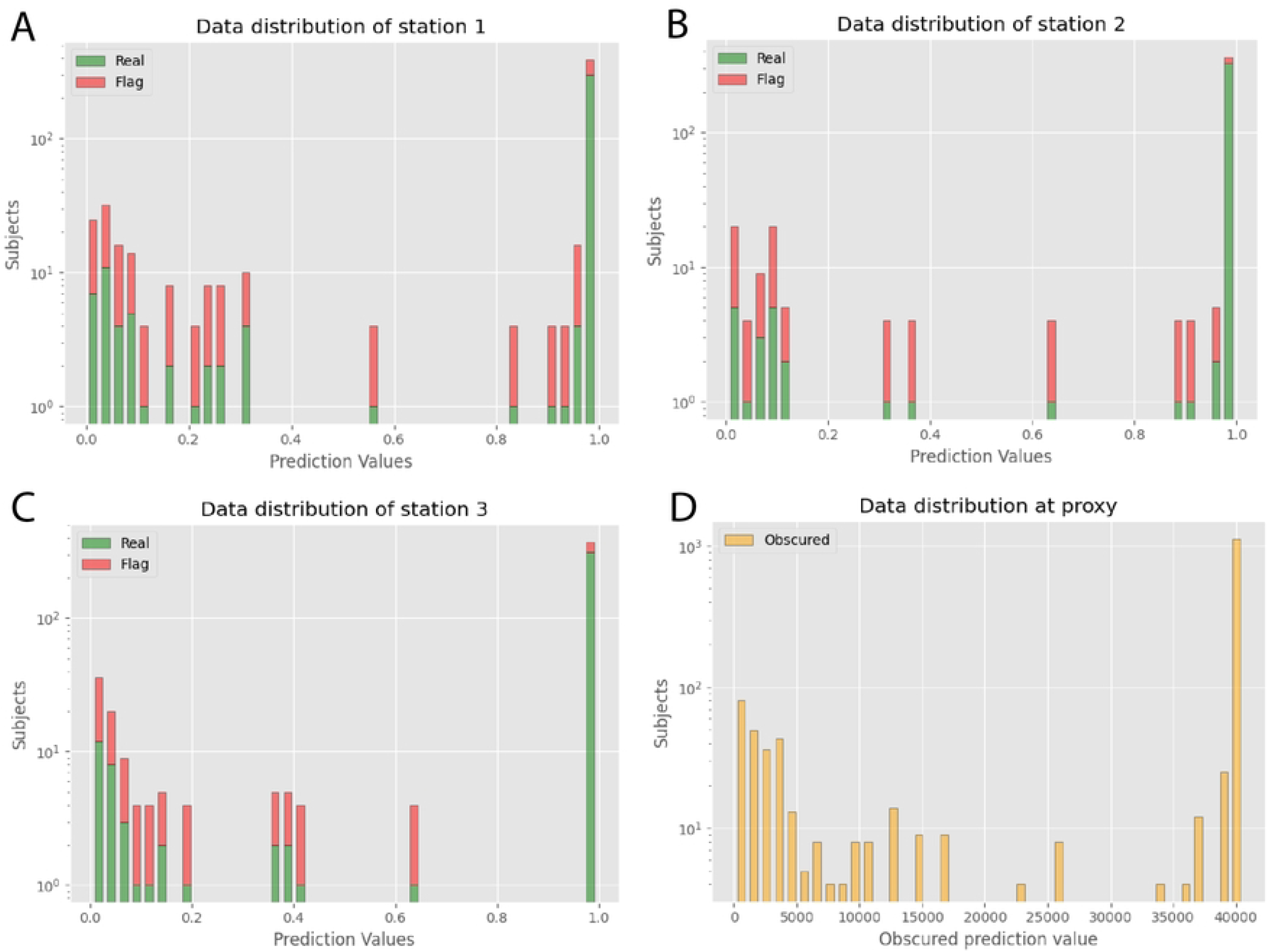
(A-C) Input data site 1-3. Prediction value distribution at Station 1 to 3, highlighting the balance between ‘Real’ and ‘Flag’ categories. Peaks at both ends of the prediction range indicate high variability in the dataset.**(D)Input data at Proxy**: Obscured prediction value distribution at the proxy station, showing transformed prediction data aggregated from all sites. The distribution indicates effective obfuscation of individual prediction values.

**Fig 3.**
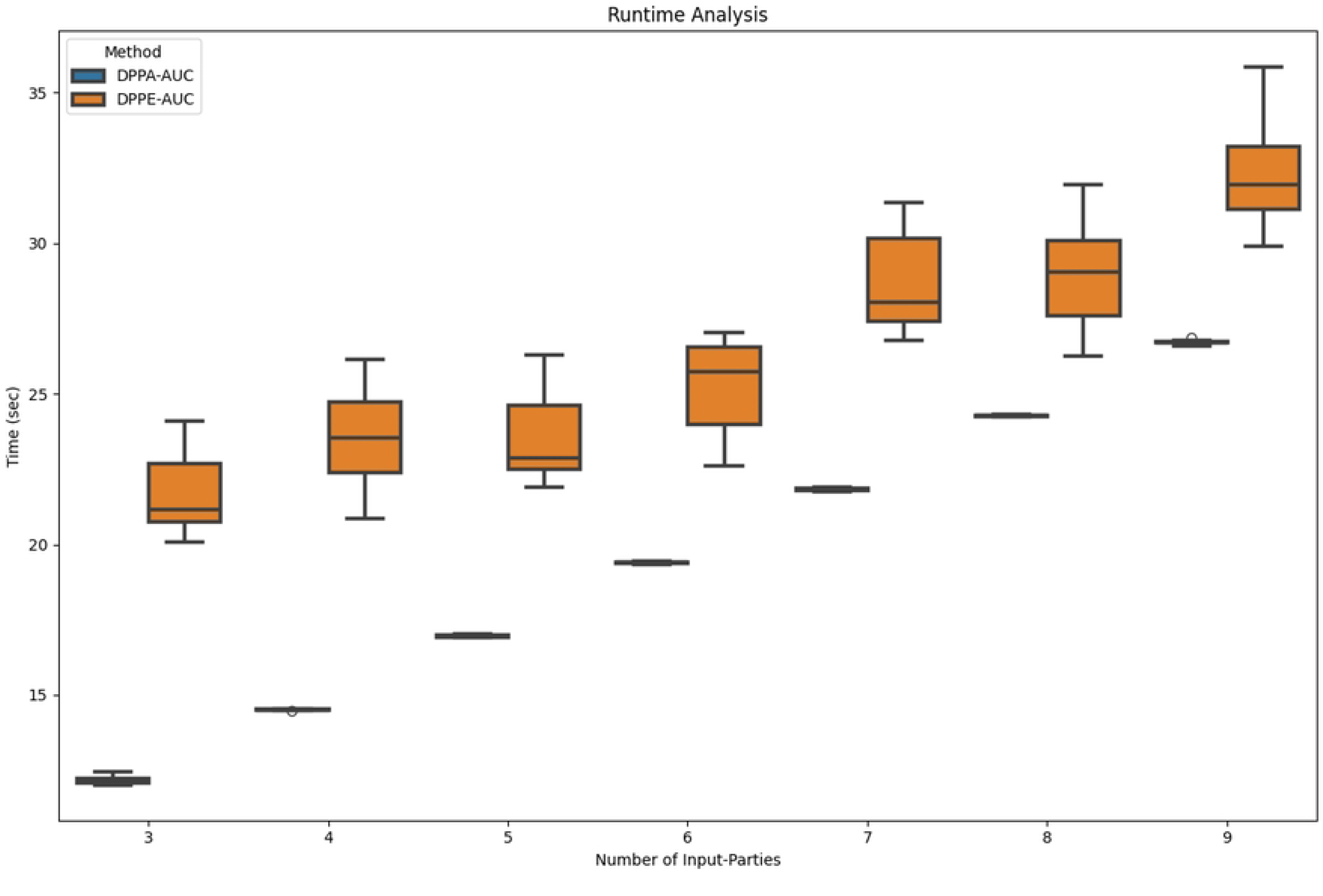
Increasing Number of Input Parties. Runtime performance over 10 runs of DPPE-AUC and DPPA-AUC methods with a fixed total dataset size of 1500 subjects. The number of participating sites (input parties) varies between 3 and 9, illustrating the scaling behavior of both methods. While execution time increased with more stations, both methods maintained robust performance.

#### Experiment 2

In this experiment, we examined how runtime performance scales with increasing sample sizes while keeping the number of stations fixed at three. This setup evaluated the computational scalability of the DPPE-AUC method compared to the DPPA-AUC method as the amount of data per station increases.

The evaluation considered up to 23,000 subjects, reflecting a practical sample size in typical PHT-meDIC studies. The results in Fig 4 demonstrated a linear increase in execution time for the DPPE-AUC method as the sample size grew. This behavior aligns with the computational demands of DPPE-AUC, where encryption and processing steps scale directly with the input data to process. In contrast, DPPA-AUC’s constant runtime offers a clear advantage in large-scale applications where efficiency and fixed computational cost are critical. The data generation followed a uniform random distribution, representing a worst-case scenario in terms of computational overhead.

**Fig 4.**
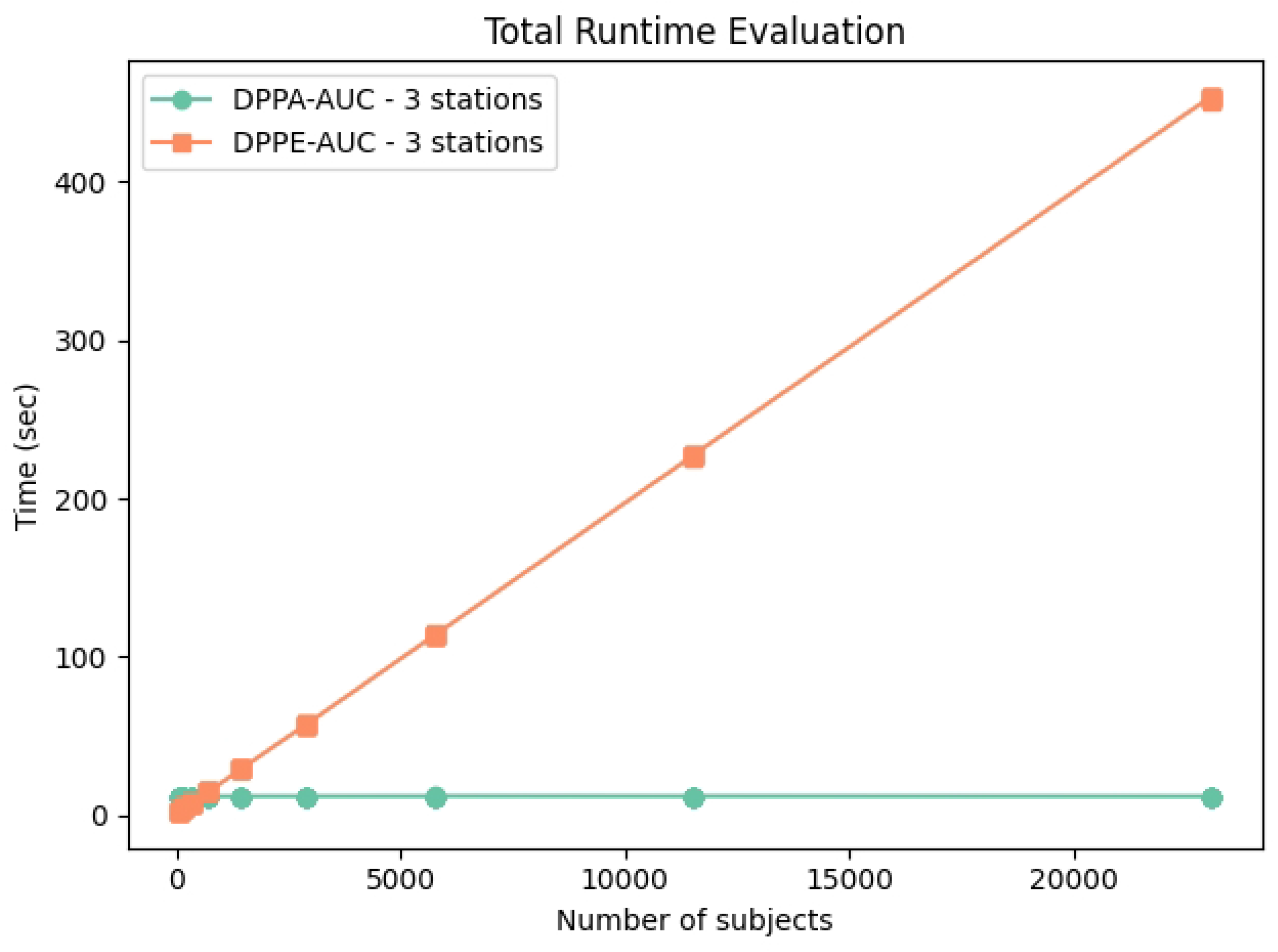
Increasing Number of Input Samples. Runtime evaluation of DPPE-AUC and DPPA-AUC methods with a fixed number of stations (3) and increasing dataset sizes (up to 23,000 subjects). DPPE-AUC exhibits linear scaling with increasing sample sizes, while DPPA-AUC maintains a constant runtime due to its reliance on a fixed number of decision points. The results emphasize DPPA-AUC’s suitability for large-scale datasets.

These findings highlight distinct scalability characteristics for the two methods. While DPPE-AUC is well-suited for moderate-scale datasets, DPPA-AUC’s fixed runtime offers a compelling advantage for large-scale applications where efficiency and predictable computational costs are critical.

## Discussion

The primary challenge identified in the current implementation for both methods is the increase in runtime as the number of participants grows, largely due to the iterative execution within the PHT-meDIC framework. To mitigate this, transitioning to a federated execution of the station-side algorithms and final computation steps would enable parallel processing, thereby significantly reducing overall total runtime, as evidenced by our experimental results.

Furthermore, the runtime performance of DPPE-AUC strongly depends on how prediction values are distributed. If every prediction value is unique, the sorting and processing steps reach their highest overhead. To both obscure the real distribution and limit uniqueness, our current approach inserts “dummy” prediction values drawn from the existing set, thereby reducing the overall number of distinct values. While this technique conceals the original distribution effectively, it remains a straightforward implementation. Consequently, there is still significant room for optimization, particularly at the proxy station during final sorting and processing, to further enhance execution speed.

The potential of achieving marginal AUC improvements, such as the ≈0.0037 difference provided by DPPE-AUC compared to the DPPA-AUC method, is mainly relevant in critical healthcare domains. In rare disease diagnoses, small gains in AUC can significantly improve early detection rates and reduce misdiagnoses [28]. Similarly, such improvements enhance early tumor detection in radiology and lower false negatives [29]. In ICU sepsis prediction, even minor performance increases can directly impact timely interventions and patient outcomes [30]. Furthermore, in personalized medicine and adverse drug reaction prediction, higher precision minimizes unnecessary treatments and associated risks [31, 32]. These examples highlight the importance of fine-grained performance gains in machine learning models for healthcare applications, emphasizing the value of DPPE-AUC.

## Conclusion

In this work, we introduced DPPE-AUC and DPPA-AUC, two cryptographic protocols that enable the computation of the global AUC in distributed and privacy-sensitive settings. DPPE-AUC calculates the AUC exactly, making it particularly useful when high-precision model assessment is important; it maintains linear scaling with the dataset size. DPPA-AUC instead relies on a sampling strategy with a fixed set of thresholds, achieving constant runtimes suitable for larger or more diverse datasets where computational efficiency is critical. Both methods are fully integrated into the PHT-meDIC framework, underscoring their practical feasibility.

By combining modified Paillier encryption, symmetric and asymmetric cryptography, and randomized encoding techniques, these protocols protect sensitive model predictions and labels while still allowing global performance metrics to be derived. This leads to a robust and flexible system that meets the privacy demands of multi-institutional collaborations in healthcare and beyond. In future work, we plan to optimize the protocols further by employing parallelization strategies, which could substantially reduce runtime in large-scale distributed networks. Through such optimizations, we aim to make secure, privacy-preserving performance evaluation an even more powerful tool for real-world medical and scientific applications.

## Data Availability

https://github.com/PHT-meDIC/PP-AUC, facilita

facilita

https://github.com/PHT-meDIC/PP-AUC

## Acknowledgments

This work has been funded by the German Federal Ministry of Health (BMG) with the project LEUKO-Expert (grant ZMVI1-2520DAT94 A-H to M.dABH). Funding from German Federal Ministry of Education and Research (BMBF) within the ‘Medical Informatics Initiative’ (DIFUTURE (01ZZ1804D), NUM-DIZ as part of “NUM 2.0”, the second funding phase of the Network of University Medicine, 01KX2121 to M.dABH and MDPPML, 01ZZ2010 to AB.U and M.A), and PrivateAIM (01ZZ2316A). M.dABH. was supported by the Tübingen AI Center.

